# Effectiveness of the 2023-2024 Formulation of the Coronavirus Disease 2019 mRNA Vaccine against the JN.1 Variant

**DOI:** 10.1101/2024.04.27.24306378

**Authors:** Nabin K. Shrestha, Patrick C. Burke, Amy S. Nowacki, Steven M. Gordon

## Abstract

**Background:** The purpose of this study was to evaluate whether the 2023-2024 formulation of the COVID-19 mRNA vaccine protects against COVID-19 caused by the JN.1 lineage of SARS-CoV-2.

**Methods:** Employees of Cleveland Clinic in employment when the JN.1 lineage of SARS-CoV2 became the dominant circulating strain, were included. Cumulative incidence of COVID-19 was examined prospectively. Protection provided by vaccination (analyzed as a time-dependent covariate) was evaluated using Cox proportional hazards regression. The analysis was adjusted for the propensity to get tested, age, sex, pandemic phase when the last prior COVID-19 episode occurred, and the number of prior vaccine doses.

**Results:** Among 47561 employees, COVID-19 occurred in 838 (1.8%) during the 16-week study period. In multivariable analysis, the 2023-2024 formula vaccinated state was associated with a significantly lower risk of COVID-19 while the JN.1 lineage was the dominant circulating strain (hazard ratio [HR], .77; 95% confidence interval [C.I.], .62-.94; *P* = .01), yielding an estimated vaccine effectiveness of 23% (95% C.I., 6%-38%). Compared to 0 or 1 prior vaccine doses, risk of COVID-19 was incrementally higher with 2 prior doses (HR, 1.46; 95% C.I., 1.12-1.90; *P* < .005), 3 prior doses (HR, 1.95; 95% C.I., 1.51-2.52; *P* < .001), and more than 3 prior doses (HR, 2.51; 95% C.I., 1.91-3.31; *P* < .001).

**Conclusions:** The 2023-2024 formula COVID-19 vaccine given to working-aged adults afforded a low level of protection against the JN.1 lineage of SARS-CoV-2, but a higher number of prior vaccine doses was associated with a higher risk of COVID-19.

**Summary:** Among 47561 working-aged Cleveland Clinic employees, the 2023-2024 formula COVID-19 vaccine was 23% effective against the JN.1 lineage of SARS-CoV-2, but a higher number of prior COVID-19 vaccine doses was associated with a higher risk of COVID-19.

## INTRODUCTION

Although the original messenger RNA (mRNA) Coronavirus Disease 2019 (COVID-19) vaccines were highly effective early in the pandemic [1,2], their effectiveness decreased as the causative agent of COVID-19, the Severe Acute Respiratory Syndrome Coronavirus 2 (SARS-CoV-2) virus, evolved over time and new variants emerged. Concurrently, the human population developed an increasing level of herd immunity, resulting in a decrease in the number of people who got infected and a substantial decline in the occurrence of severe illness among those who got infected.

Newer vaccines were developed in an attempt to overcome diminishing vaccine effectiveness. Bivalent COVID-19 mRNA vaccines, which encoded antigens represented in the original vaccine as well as antigens representing the BA.4/5 lineages of the Omicron variant, were approved by the United States Food and Drug Administration (FDA), in August 2022. In our cohort of healthcare personnel in northern Ohio, we found that the effectiveness of the bivalent vaccines against COVID-19 decreased from 29% when the Omicron BA.4/5 lineages were the predominant circulating strains, to 19% when the BQ lineages were dominant, to no longer effective by the time the XBB lineages became dominant [3].

Moderna TX Inc. and Pfizer-BioNTech Inc. updated their mRNA COVID-19 vaccines (the updated ones designated as the 2023-2024 formulation) to more closely target circulating variants. Both of these vaccines encoded the spike protein of SARS-CoV-2 Omicron variant lineage XBB.1.5 (Omicron XBB.1.5) [4]. These updated vaccines were approved for emergency use by the FDA on 11 September 2023 [5], and the next day the CDC recommended them for everyone 6 months and older [6]. However, by the time these vaccines became available to the public the XBB lineages were already no longer the dominant circulating strains in many parts of the USA, having been supplanted by the HV.1, EG.5, and other lineages [7]. Despite this, the 2023-2024 formulation of the COVID-19 vaccine had an effectiveness around 42% in the early weeks after the vaccine became available [8], providing reassurance that the vaccine was still effective even when the strain the vaccine targeted was no longer the dominant circulating strain. However, our earlier study only included a short duration while the JN.1 lineage was dominant, and was not able to show a statistically significant protective effect after the JN.1 lineage became the dominant circulating strain [8]. It is possible that the small number of events over the short duration of follow-up during this phase may have resulted in a protective effect being missed.

The purpose of this study was to evaluate, in a study with longer follow-up, whether the 2023-2024 formula COVID-19 vaccine protects against COVID-19 at a time when the JN.1 lineage of SARS-CoV2 is the predominant circulating strain.

## METHODS

### Study design

This was a prospective cohort study conducted at the Cleveland Clinic Health System (CCHS) in the United States.

### Patient Consent Statement

The study was approved by the Cleveland Clinic Institutional Review Board as exempt research (IRB no. 22-917). A waiver of informed consent and waiver of HIPAA authorization were approved to allow the research team access to the required data.

### Setting

Cleveland Clinic has always given very high priority to employee access to COVID-19 testing and COVID-19 vaccination and has carefully studied the effectiveness of the various iterations of the COVID-19 vaccines [9–11,3,12,8]. The JN.1 lineage of SARS-CoV-2 emerged near the end of the year 2023, and became the predominant circulating strain in Ohio around 31 December 2023. This date was considered this study’s start date.

### Participants

CCHS employees in employment at any Cleveland Clinic location in Ohio on the study start date were included in the study. Those for whom age and gender were not available were excluded.

### Variables

Covariates collected were age, sex, and job location, as described in our earlier studies [9,11,10,3,12,8]. Institutional data governance rules related to employee data limited our ability to supplement our dataset with additional clinical variables. Subjects were considered pre-pandemic hires if hired before 16 March 2020, the day COVID-19 testing became available in our institution, and pandemic hires if hired on or after that date.

Number of prior vaccine doses was the number of doses of older COVID-19 vaccines received. Prior COVID-19 was defined as a positive nucleic acid amplification test (NAAT) for SARS-CoV-2 any time before the study start date. The date of infection for a prior episode of COVID-19 was the date of the first positive test for that episode of illness. A positive test more than 90 days following the date of a previous infection was considered a new episode of infection. The propensity to get tested for COVID-19 was defined as the number of COVID-19 NAATs done divided by the number of years of employment at CCHS during the pandemic, before the study start date.

The distribution of circulating variants in Ohio at any time was obtained from monitoring data from the CDC [7]. The pandemic phase for prior episodes of COVID-19 (pre-Omicron, pre-XBB Omicron, and XBB Omicron or later) was defined by which variant/lineages accounted for more than 50% of infections in Ohio at the time of the infection [7].

### Outcome

The study outcome was time to COVID-19, the latter defined as a positive NAAT for SARS-CoV-2 any time after the study start date. Outcomes were followed until 22 April 2024, allowing for evaluation of outcomes up to 16 weeks from the study start date.

### Statistical analysis

Subjects infected with COVID-19 in the weeks preceding the study start date were not included until 90 days had passed since their recent prior infection. Individuals were considered vaccinated 7 days after receipt of a single dose of the 2023-2024 formula COVID-19 vaccine. Subjects whose employment was terminated during the study period before they had COVID-19 were censored on the date of termination.

A Simon-Makuch hazard plot [13] was created to compare the cumulative incidence of COVID-19 in the vaccinated and non-vaccinated states with respect to the 2023-2024 formula COVID-19 vaccine, by treating such vaccination as a time-dependent covariate. Curves for the non-vaccinated state were based on data while the vaccination status of subjects, with respect to the 2023-2024 formula COVID-19 vaccine, remained “non-vaccinated”. Curves for the vaccinated state were based on data from the date the vaccination status changed to “vaccinated”.

A Cox proportional hazards regression model was fit to examine the association of various variables with time to COVID-19. Vaccination with the 2023-2024 formula COVID-19 vaccine was included as a time-dependent covariate whose value changed from “non-vaccinated” to “vaccinated” 7 days after receipt of the vaccine [14]. The possibility of multicollinearity in the models was evaluated using variance inflation factors. The proportional hazards assumption was checked using log(-log(survival)) vs. time plots. Vaccine effectiveness (VE) was calculated from the hazard ratios (HRs) for 2023-2024 formula COVID-19 vaccination in the multivariable model using the formula VE = 1 - HR.

The analysis was performed by N. K. S. and A. S. N. using the *survival* package and R version 4.2.2 [14–16].

## RESULTS

Of the 47 561 employees included in the study, 7598 had received the 2023-2024 formula COVID-19 vaccine before the study start date. By the end of the study 8613 (18%) had received the 2023-2024 formula COVID-19 vaccine, which was the Pfizer vaccine in 7589 (89%). One hundred and seven subjects (0.2%) were censored during the study because of termination of employment. Altogether, 838 employees (1.8%) acquired COVID-19 during the 16 weeks of the study.

### Baseline characteristics

Table 1 shows the characteristics of subjects included in the study. The mean age of study subjects was 42 years, and almost 75% were female. Among these, 22 389 (47%) had previously had a documented episode of COVID-19 and 17 630 (37%) had previously had an Omicron variant infection. 41 387 subjects (87%) had previously received at least one dose of a COVID-19 vaccine, 39 552 (83%) had received at least two doses, and 43 002 (90%) had been previously exposed to SARS-CoV-2 by infection or vaccination.

**Table 1.** Baseline characteristics of the 47561 employees of Cleveland Clinic in Ohio included in the study.

| **Characteristics** | **All subjects**<br>**(n = 47561)** |
| --- | --- |
| **Age in years, mean (SD)** | 42.0 (13.4) |
| **Sex** |  |
| Female | 35 436 (74.5) |
| Male | 12 125 (25.5) |
| **Location** |  |
| Cleveland Clinic Main | 19 712 (41.4) |
| Cleveland area regional hospitals | 12 087 (25. 4) |
| Ambulatory centers | 8830 (18. 6) |
| Cleveland Clinic Akron | 3992 (8.4) |
| Administrative centers | 1765 (3.7) |
| Cleveland Clinic Medina | 1175 (2.5) |
| **Hire cohort** |  |
| Pre-pandemic | 27 802 (58.5) |
| Pandemic | 19 759 (41.5) |
| **Pandemic phase during which most recent infection occurred** |  |
| Not previously known to be infected | 25 172 (52.9) |
| Pre-Omicron | 4759 (10.0) |
| Omicron pre-XBB | 12 723 (26.8) |
| Omicron XBB or later | 4907 (10.3) |
| **Days since most recent infection, mean (SD)** | 577 (317) |
| **Prior vaccination history** |  |
| **Monovalent and bivalent vaccines** | 12 173 (25.6) |
| **Bivalent vaccine only** | 376 (<1) |
| **Monovalent vaccine only** | 27 948 (58.8) |
| **Non-mRNA vaccine only** | 890 (1.9) |
| **No vaccine** | 6174 (13.0) |
| **Number of prior vaccine doses** |  |
| **0** | 6174 (13.0) |
| **1** | 1835 (3.9) |
| **2** | 13 232 (27.8) |
| **3** | 14 147 (29.7) |
| **>3** | 12 173 (25.6) |
| **Days since most recent vaccine, mean (SD)** | 688 (219) |
| **Days since proximate SARS-CoV-2 exposure<sup>a</sup>, mean (SD)** | 579 (253) |

Data are presented as no. (%) unless otherwise indicated.
<sup>a</sup>By infection or vaccination

### Effectiveness of the 2023-2024 formula COVID-19 mRNA vaccine

There was no significant difference in the cumulative incidence of COVID-19 in the 2023-2024 formula vaccinated state compared to the non-vaccinated state in an unadjusted analysis (Figure 1).

**Figure 1.**
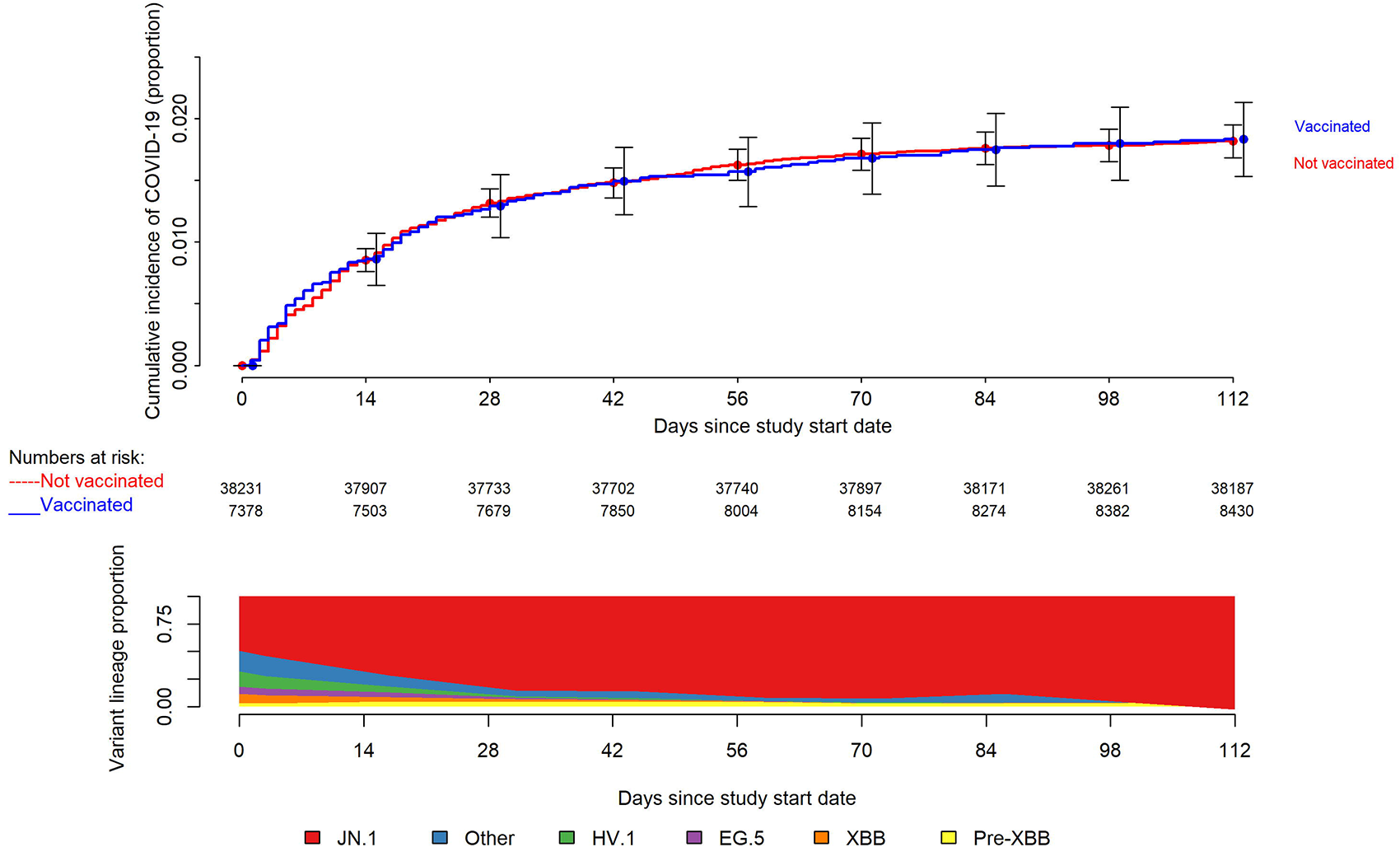
Simon-Makuch plot comparing the cumulative incidence of COVID-19 for the vaccinated and non-vaccinated states with respect to the 2023-2024 formulation COVID-19 vaccine. Day zero was 31 December 2023, the day the JN.1 lineage of SARS-CoV-2 was projected to have become the dominant circulating strain in Ohio. Individuals with recent past infections do not contribute to the denominator until it has been at least 90 days since their prior infection. Point estimates and 95% confidence intervals are jittered along the x-axis to improve visibility. Variant proportions are based on data from the CDC, considered to be in the middle of the week reported, grouped into pre-XBB, XBB, HV.1, EG.5, JN.1, and other lineages, and presented as an area plot with values extrapolated for day zero and the study end date using linear regression models with data for the weeks immediately before and after the target date.

In a multivariable Cox proportional hazards regression model, adjusted for propensity to get tested for COVID-19, age, sex, hire cohort, number of prior COVID-19 vaccine doses, and epidemic phase when the last prior COVID-19 episode occurred, vaccination with the 2023-2024 formula COVID-19 vaccine provided some protection against COVID-19 in the first 16 weeks after the JN.1 lineage of SARS-CoV2 became the dominant strain in Ohio (hazard ratio [HR], .77; 95% confidence interval [C.I.], .62-.94; *P* = 0.01). Point estimates and 95% confidence intervals for hazard ratios for the variables included in the unadjusted and adjusted Cox proportional hazards regression models are shown in Table 2. The calculated overall vaccine effectiveness from the model was 23% (95% C.I., 6%-38%).

**Table 2.** Unadjusted and Adjusted Associations with Time to COVID-19.

| Variables | Unadjusted HR<br>(95% CI) | P | Adjusted HR<sup>a</sup><br>(95% CI) | P |
| --- | --- | --- | --- | --- |
| **2023-2024 formulation vaccinated state<sup>b</sup>** | 1.01 (.84-1.21) | .92 | .77 (.62-.94) | 0.01 |
| **Propensity to get tested for COVID-19** | 1.04 (1.03-1.05) | <.001 | 1.04 (1.03-1.05) | <.001 |
| **Age** | 1.005 (1.000-1.010) | .06 | 1.001 (.995-1.006) | .86 |
| **Male sex<sup>c</sup>** | .78 (.66-.92) | .003 | .79 (.67-.93) | .006 |
| **Pre-pandemic hire<sup>d</sup>** | 1.20 (1.04-1.38) | .01 | .99 (.84-1.16) | .92 |
| **Last prior infection phase (ref: pre-Omicron infection)** |  |  |  |  |
| **No prior documented infection** | .78 (.62-.97) | .03 | .80 (.64-1.01) | 0.06 |
| **Omicron Pre-XBB** | 1.17 (.93-1.47) | .19 | 1.08 (.86-1.36) | .51 |
| **Omicron XBB or later** | .66 (.47-.94) | .03 | .59 (.41-.84) | .004 |
| **Number of prior vaccine doses (ref: 0 or 1)** |  |  |  |  |
| **2** | 1.58 (1.22-2.05) | <.001 | 1.46 (1.12-1.90) | .005 |
| **3** | 2.03 (1.58-2.60) | <.001 | 1.95 (1.51-2.52) | <.001 |
| **>3** | 2.29 (1.79-2.94) | <.001 | 2.51 (1.91-3.31) | <.001 |

Abbreviation: COVID-19, Coronavirus Disease 2019; HR, hazard ratio; CI, confidence interval;
<sup>a</sup>From a multivariable Cox-proportional hazards regression model with the 2023-2024 formulation vaccinated state treated as a time-dependent covariate.
<sup>b</sup>Time-dependent covariate
<sup>c</sup>Reference is female sex
<sup>d</sup>Reference is pandemic hire

The multivariable analysis also found that compared to receipt of 0 or 1 prior vaccine doses, risk of COVID-19 was increasingly higher with receipt of 2 prior doses (HR, 1.46; 95% C.I., 1.12-1.90; *P* < .005), 3 prior doses (HR, 1.95; 95% C.I., 1.51-2.52; *P* < .001), and more than 3 prior doses (HR, 2.51; 95% C.I., 1.91-3.31; *P* < .001). If number of prior vaccine doses was not adjusted for in the multivariable model, the 2023-2024 formulation of the vaccine was not protective against COVID-19 (HR 1.01, 95% C.I. .84 – 1.21, *P* = 0.95).

## DISCUSSION

This study found that the 2023-2024 formula COVID-19 mRNA vaccine was about 23% effective overall in protecting against infection with the JN.1 lineage of SARS-CoV-2. Although the vaccine was designed to target the XBB lineages of the Omicron variant, the vaccine was notably still effective even though practically all the infections occurring in the community during the study period were caused by the JN.1 lineage of the virus.

The strengths of our study include its reasonably large sample size and a robust methodology that has been refined by reviews over multiple publications [3,8–12]. Vaccine effectiveness is calculated from risk differences and should provide more accurate estimates than by extrapolations from odds ratios obtained from case-control studies as is commonly done [17]. Treating vaccination with the 2023-2024 formulation of the COVID-19 vaccine as a time-dependent covariate allows for determining vaccine effectiveness in real time, thereby providing results in a time frame when they are still meaningful. Adjusting for hire cohort (pre-pandemic versus pandemic) in the multivariable analysis would have mitigated against bias that might arise from possible incomplete information on prior infection and prior vaccination among the pandemic hires compared to the pre-pandemic hires. The potential for risk of bias from difference in testing for COVID-19 among individuals inclined and not inclined to get vaccinated was mitigated by adjusting for the propensity of an individual to get tested for COVID-19.

The study has several limitations, which have all been discussed extensively in prior publications [3,10-12]. Individuals with unrecognized prior infection would have been misclassified as previously uninfected. There could be concern that such misclassification could result in underestimating the protective effect of the vaccine, because the protection against disease afforded by natural immunity from prior infection would limit the detection of a vaccine effect. However, such misclassification would have occurred in both vaccinated and non-vaccinated states, and there is little reason to suppose that prior infections would have been missing in the vaccinated and nonvaccinated states at rates disproportionate enough to significantly affect the results of the study. The widespread availability of home testing kits might have reduced detection of incident infections, but there is little reason to suppose a significant difference in home testing between the vaccinated and unvaccinated states. This potential effect should also be somewhat mitigated in our healthcare cohort because one needs a NAAT to get paid time off, providing a strong incentive to get a NAAT if one tests positive at home. We were unable to distinguish between symptomatic and asymptomatic infections. The number of severe illnesses was too small to examine as an outcome. Like our previous studies, this study was done in a healthcare population, and included no children and few elderly subjects, and most subjects would not have been expected to be immunocompromised.

Consistent with similar findings in many prior studies [3,8,10,12,18–20], a higher number of prior vaccine doses was associated with a higher risk of COVID-19. The exact reason for this finding is not clear. It is possible that this may be related to the fact that vaccine-induced immunity is weaker and less durable than natural immunity. So, although somewhat protective in the short term, vaccination may increase risk of future infection because the act of vaccination prevents the occurrence of a more immunogenic event. Thus, the short-term protection provided by a COVID-19 vaccine comes with a risk of increased susceptibility to COVID-19 in the future. This understanding suggests that a more nuanced approach to COVID-19 is necessary. Although some individuals are at high risk of complications from COVID-19, and may benefit from receiving a vaccine frequently, the wisdom of vaccinating everyone with a vaccine of low effectiveness every few months to prevent what is generally a mild or an asymptomatic infection in most healthy persons, needs to be questioned.

In conclusion, this study found an overall low protective effect of the 2023-2024 formula COVID-19 vaccine against infection with the JN.1 lineage of SARS-CoV-2, while also finding that a higher number of prior vaccine doses was associated with a higher risk of COVID-19.

## Notes

### Author contributions

N. K. S.: Conceptualization, methodology, validation, investigation, data curation, software, formal analysis, visualization, writing-original draft preparation, writing-reviewing and editing, supervision, project administration. P. C. B.: Resources, investigation, validation, writing-reviewing and editing. A. S. N.: Methodology, formal analysis, visualization, validation, writing-reviewing and editing. S. M. G.: Project administration, resources, writing-reviewing and editing.

## Data Availability

All data produced in the present study are available upon reasonable request to the authors

